# Exploring Attitudes and Acceptance of Artificial Intelligence in Multiple Sclerosis from the Patient Perspective

**DOI:** 10.64898/2026.01.27.26344998

**Authors:** Hernan Inojosa, Lars Masanneck, Isabel Voigt, Dirk Schriefer, Nele von Horsten, Judith Wenk, Iva Gasparovic-Curtini, Rocco Haase, Sven Meuth, Hagen B. Huttner, Stephen Gilbert, Marc Pawlitzki, Tjalf Ziemssen

## Abstract

Artificial intelligence (AI) is increasingly being integrated into healthcare, particularly in data-intensive chronic diseases that rely on longitudinal monitoring and shared decision-making. Multiple sclerosis is a prototypical example of such care, but real-world benefit will depend on whether people accept AI support in different clinical roles. We conducted a cross-sectional, web-based survey among 241 people with MS (pwMS) to assess comfort with AI across eight clinical domains and to identify predictors of acceptance. We derived an artificial-intelligence attitudes composite with high internal consistency (Cronbach alpha = 0.90). Overall acceptance was moderate (mean 3.39 ± 0.78). Acceptance differed across domains, demonstrating a responsibility gradient: comfort was highest for supportive applications such as chronic management (54.4%) and symptom screening (50.2%), but lower for treatment selection (38.6%) and diagnosis (35.3%; P < 0.001). In multivariable models, frequent general AI use (at least weekly; 30.7%) was the strongest independent predictor of acceptance (P < 0.001). Acceptance also differed by region (Eastern vs Western Germany, P = 0.025), whereas clinical disability was not significantly associated. Older age was associated with lower acceptance of AI-supported management. Most participants viewed AI as a logistical support tool but, assuming equal diagnostic accuracy, 78.8% preferred joint artificial-intelligence-clinician decision-making with clinician final responsibility. These findings indicate that acceptance is context-dependent and aligns more closely with prior familiarity than with disease severity. Implementation should move beyond technical validation to transparent, clinician-led’human-in-the-loop’ workflows with explicit accountability and staged adoption beginning with low-risk use cases.

**Author Summary:** We use artificial intelligence more and more in everyday life, and similar tools are now being introduced into medical care. For long-term conditions such as multiple sclerosis, digital systems could help manage large amounts of clinical information and support monitoring between visits. At the same time, these tools will only be useful if the people receiving care are willing to use them and understand what role they play.

In this study, we asked 241 people living with multiple sclerosis in Germany how comfortable they would feel with artificial intelligence in different parts of care. We found that comfort depended strongly on the task. Participants were most open to artificial intelligence when it supported practical, lower-risk functions such as ongoing monitoring or symptom screening, and they were more cautious when it was described as influencing diagnosis or treatment choices. Most participants wanted clinicians to remain responsible for final decisions. Acceptance was higher among people who already used artificial intelligence frequently in everyday life, and it differed by age and by region. Our findings suggest that successful implementation will require more than technical performance: it should be introduced transparently, with clinician oversight, and in a stepwise way that builds familiarity without shifting responsibility away from the clinical team.

## Introduction

Artificial intelligence (AI) is rapidly entering clinical workflows, promising to transform the management of complex chronic conditions. Multiple sclerosis (MS) serves as a critical model for this transition: as a paradigm of data-driven care, AI is expected to influence imaging, risk stratification, prognosis, and long-term monitoring (1). However, the clinical value of AI will translate into routine care only if patients consider these systems acceptable and trustworthy. This is particularly relevant in MS, where many AI-enabled services depend on continuous patient-generated data (e.g., digital biomarkers, patient-reported outcomes) and are intended to support shared decision-making rather than replace clinician judgement (2–5).

People with MS (pwMS) represent a population with comparatively high digital engagement, which may facilitate adoption of AI-supported care models (6). Earlier work has shown frequent internet use and widespread uptake of digital communication among pwMS, and acceptance of newer communication channels was higher among those already using established technologies (4, 7, 8). This baseline digital engagement provides a lever for introducing AI-based tools tailored to this population. Given the demographic profile of MS, which disproportionately affects females and typically begins in early adulthood, this population may further support digital readiness and openness toward AI-driven health innovations (9, 10).

While digital literacy is high, acceptance of automated and AI-based systems in healthcare is not universal (4, 11, 12). Beyond individual familiarity, acceptance is also shaped by trust in the health system itself (13). This creates an opportunity to introduce AI applications that build on existing digital behaviours. Yet, digital access does not automatically imply readiness to rely on algorithmic decision support. Evidence from other healthcare settings suggests that acceptance of automated or AI-based systems is heterogeneous and shaped by sociodemographic context, technology familiarity, and perceived risk (4, 11, 14). For example, studies in primary care and hospital populations indicate that trust in AI varies across groups and tends to be higher among individuals with greater AI knowledge and more frequent technology use, while some patients - particularly those who feel vulnerable, express reservations about automation in high-stakes decisions (4, 11) In chronic disease contexts, patients often prefer AI as an assistive “support tool,” valuing convenience and monitoring while maintaining a strong preference for human accountability in critical decisions (5).

In the context of MS, recent perspectives emphasize that successful AI implementation requires patient input and co-design to address concerns around transparency, privacy, and responsibility (15, 16). Despite this, empirical data describing how pwMS perceive AI, both in general and across distinct clinical use cases, remain scarce. Given the heterogeneity of the MS population (age, disability, socioeconomic background) and the long-term nature of care, understanding where pwMS are comfortable with AI and which factors predict acceptance is essential for responsible and effective implementation (16).

Therefore, this study investigated attitudes toward AI among pwMS in Germany, distinguishing general acceptance from comfort with specific clinical domains and examining how demographic, clinical, and AI-exposure variables relate to these perceptions.

## Results

### Study Population and Clinical Characteristics

A total of 241 participants were included. Participant characteristics are shown in Table 1. Most respondents were female (73.5%), had relapsing–remitting MS (61.4%), and reported low-to-moderate disability (PDDS median 2.0 [IQR 1.0–4.0]). Mean age was 50±11 years and mean disease duration 13.5±9.8 years. Age correlated positively with disability (r_s_ =0.434, P < 0.001) and disease duration (r_s_ = 0.418, P < 0.001); disease duration was also positively associated with PDDS (r_s_ = 0.359, P < 0.001).

**Table 1.**
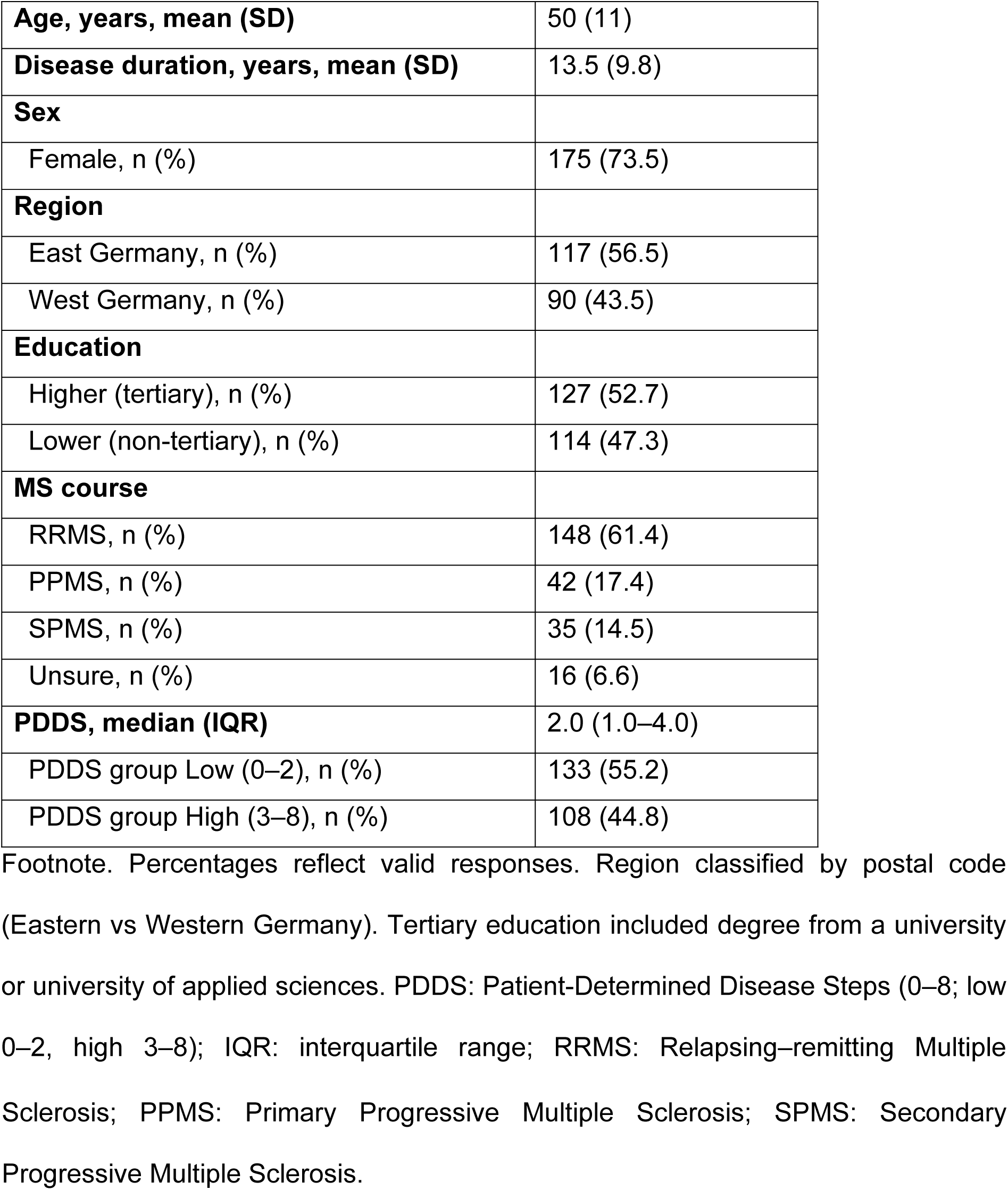
Baseline demographic and clinical characteristics (N=241)

### Digital Engagement and Technological Literacy

Technology access was high: 95.8% reported access to a smartphone/tablet. Daily use was common (smartphone ≥1/day 99.6%, internet ≥1/day 94.6%, computer ≥1/day 75.9%). Among wearable users (n=83), 88.0% reported daily use (Supplementary Material 2). Devices were frequently used for MS-related information seeking (computer/laptop 58.3%; smartphone/tablet 67.7%; Supplementary Material 3).

### AI literacy, exposure, and attitudes across domains

Nearly half of respondents rated their AI knowledge as good/expert (47.7%). General AI use ≥1×/week was reported by 30.7%, whereas health-related AI use ≥1×/week was less common (10.8%; Supplementary Material 4).

Acceptance differed significantly across the eight assessed MS-care domains (Cochran’s Q(7)=98.89, P < 0.001; Fig 1), suggesting a “responsibility gradient.” Respondents were largely optimistic about AI’s potential (60.5% somewhat/very optimistic) and supported increasing AI use in MS care (56.9% agree/strongly agree). Comfort was highest for supportive applications, including chronic management (54.4%) and symptom screening (50.2%), but decreased for high-stakes use cases, notably treatment selection (38.6%) and diagnosis (35.3%).

**Fig 1.**
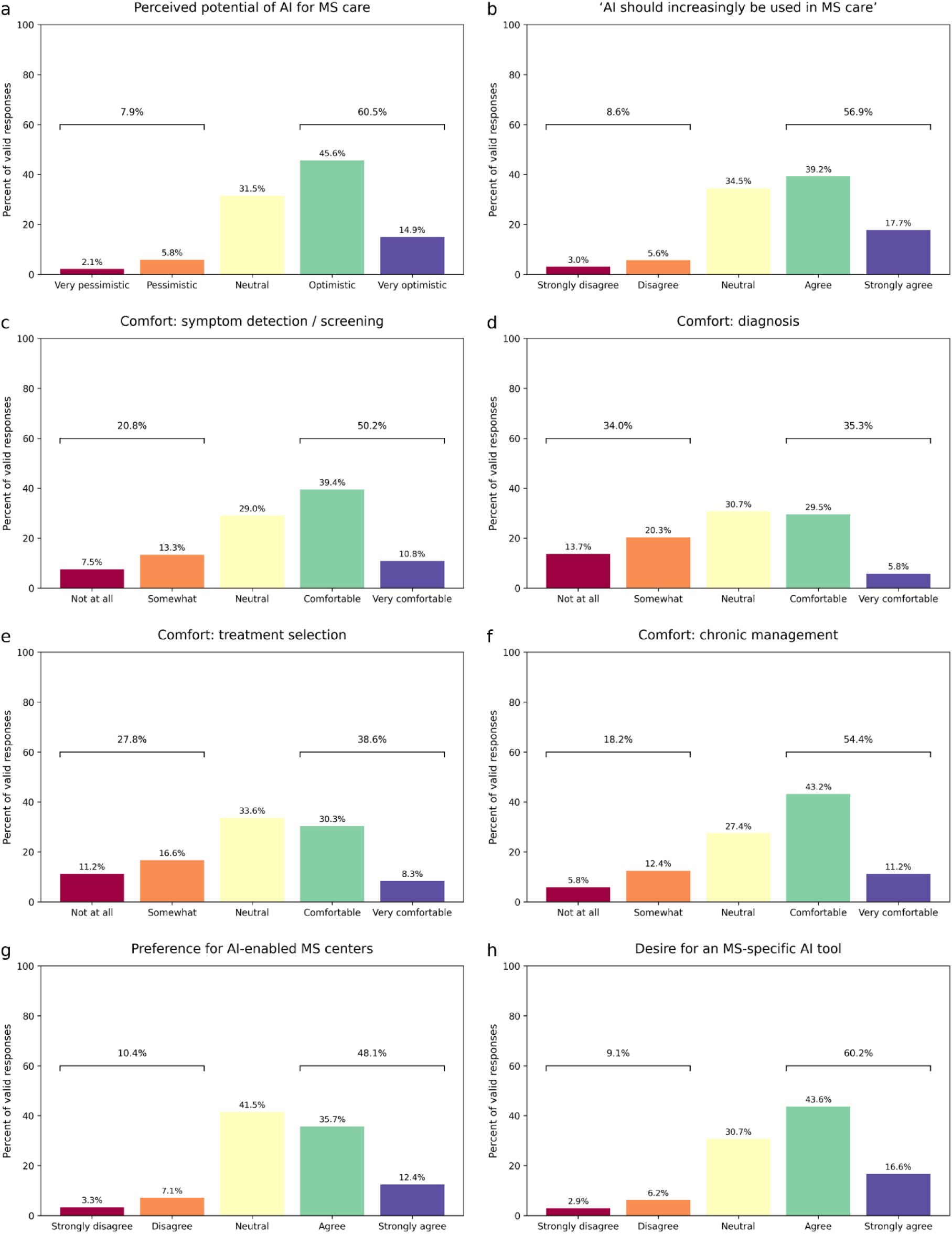
Attitudes toward AI in MS care (Likert distributions, % of valid responses). Five-point Likert distributions shown as percent of valid responses (N=241). Panels: (a) perceived potential; (b) agreement with increased use; (c-f) comfort across use-cases (symptom detection, diagnosis, treatment selection, chronic management); (g) preference for AI-enabled centres; (h) desire for an MS-specific tool. Responses skewed toward optimism and comfort for management/screening, with more neutral or mixed views for diagnosis/treatment. AI: Artificial Intelligence. MS: Multiple Sclerosis.

**Fig 2.**
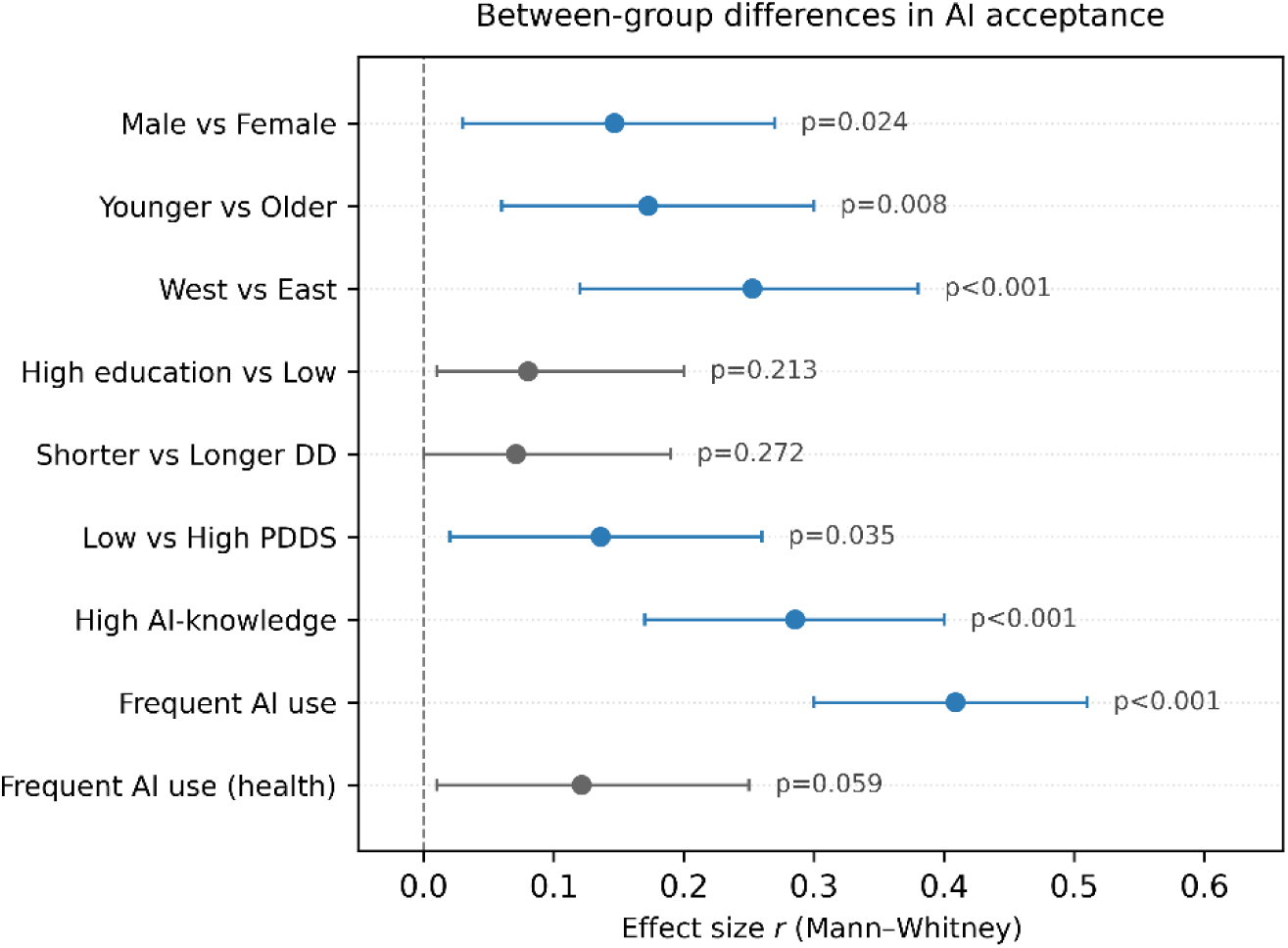
Between-group differences in AI acceptance (composite). Forest plot of Mann–Whitney effect sizes (*r*) for between-group differences in the AI-attitudes composite score across predefined subgroups. Points represent effect sizes; horizontal lines indicate 95% confidence intervals. Larger absolute values indicate greater between-group differences. The sign of *r* reflects direction: positive values indicate higher AI-attitudes in the first group named on the y-axis. Blue points denote P < 0.05 (two-tailed); grey points denote non-significant comparisons. Annotated labels show exact p-values. AI: artificial intelligence; DD: disease duration; PDDS: Patient-Determined Disease Steps.

Regarding adoption preferences, 48.1% agreed they would prefer treatment at an AI-enabled MS centre, and 60.2% expressed interest in an MS-specific AI tool to be used alongside their clinician. When asked about diagnostic decisions assuming equal accuracy, the majority preferred joint AI–clinician decision-making with clinician final responsibility (78.8%); 7.9% preferred clinician-only decisions and 0.8% preferred AI as the final decision-maker.

### Overall acceptance (AI attitudes composite) and subgroup differences

The AI attitudes composite showed excellent internal consistency (Cronbach’s α = 0.90). Overall acceptance was moderate (mean 3.39±0.78; median 3.50 [IQR 3.00–3.88]). In unadjusted analyses, higher composite scores were observed in men (P = 0.024), younger participants (P = 0.008), those living in Western vs Eastern Germany (P < 0.001), participants with lower disability (PDDS 0–2 vs ≥3; P = 0.035), those with higher AI knowledge (P < 0.001), and those reporting frequent general AI use (≥1×/week; P < 0.001). Differences by education, disease duration, and health-related AI use were not statistically significant (P = 0.213, 0.272, and 0.059, respectively; Supplementary Material 5).

### Independent Predictors of AI Acceptance

In the multivariable linear regression model for the AI attitudes composite (Table 2), the model explained 26% of variance (adj. R²=0.226; F(9,194)=7.57; P < 0.001; VIF≤1.71). Frequent general AI use emerged as the strongest independent predictor (B=0.58, 95% CI 0.34–0.82; P < 0.001), followed by residence in Western Germany (B=0.21, 95% CI 0.03–0.39; P = 0.025). Age, education, gender, disease duration, PDDS, AI knowledge, and health-related AI use were not independently associated with the composite (Supplementary material 6).

**Table 2.** Predictors of AI acceptance (AI attitudes composite)

| Predictor | B | 95% CI for B | $\beta$ (std.) | P |
| --- | --- | --- | --- | --- |
| Education (High vs Low) | 0.074 | -0.110 to 0.258 | 0.052 | 0.429 |
| Age (years) | -0.003 | -0.013 to 0.007 | -0.052 | 0.489 |
| Gender (Male vs Female) | -0.042 | -0.256 to 0.172 | -0.026 | 0.702 |
| Region (West vs East) | 0.211 | 0.029 to 0.393 | 0.146 | **0.025** |
| Disease duration (years) | 0.002 | -0.008 to 0.012 | 0.032 | 0.665 |
| Disability (PDDS) | -0.036 | -0.083 to 0.011 | -0.109 | 0.126 |
| AI knowledge (High vs Low) | 0.070 | -0.132 to 0.272 | 0.049 | 0.500 |
| General AI use (Frequent $\geq 1 \times / \text{week}$ vs Infrequent $< 1 \times / \text{week}$) | 0.581 | 0.338 to 0.824 | 0.379 | **<0.001** |
| Health-related AI use (Frequent vs Infrequent) | 0.070 | 0.251 to 0.391 | 0.030 | 0.669 |

Footnote: Results from a multivariable linear regression model with the AI attitudes composite score as the dependent variable ( $R^2=0.29$ ). B: unstandardized regression coefficient; 95%CI: 95% confidence interval for B; $\beta$ (std.): standardized regression coefficient. AI: Artificial Intelligence; PDDS: Patient Determined Disease Steps. Reference categories for categorical predictors are: Education (Low), Gender (Female), Region (East), AI knowledge (Low), General AI use (Infrequent), and Health-related AI use (Infrequent). Significant predictors ( $P < 0.05$ ) are highlighted in bold.

Across proportional-odds models for individual AI-preference outcomes, frequent general AI use was the most consistent predictor of greater acceptance, including optimism about AI, support for increased deployment, comfort with AI for symptom detection, treatment selection, and chronic disease management, preference for AI-enabled centres, and interest in an MS-specific AI tool. Western German residence was associated with higher acceptance for perceived potential for AI-supported, chronic management, preference for AI-enabled centres, and desire for an MS-specific tool. Higher AI knowledge was associated with greater comfort for screening only, and older age was associated with lower comfort for AI-supported chronic management. Notably, comfort with AI for diagnosis showed a regional association (lower in the East), whereas general AI use was not a significant predictor in that domain (Fig 3).

**Fig 3.**
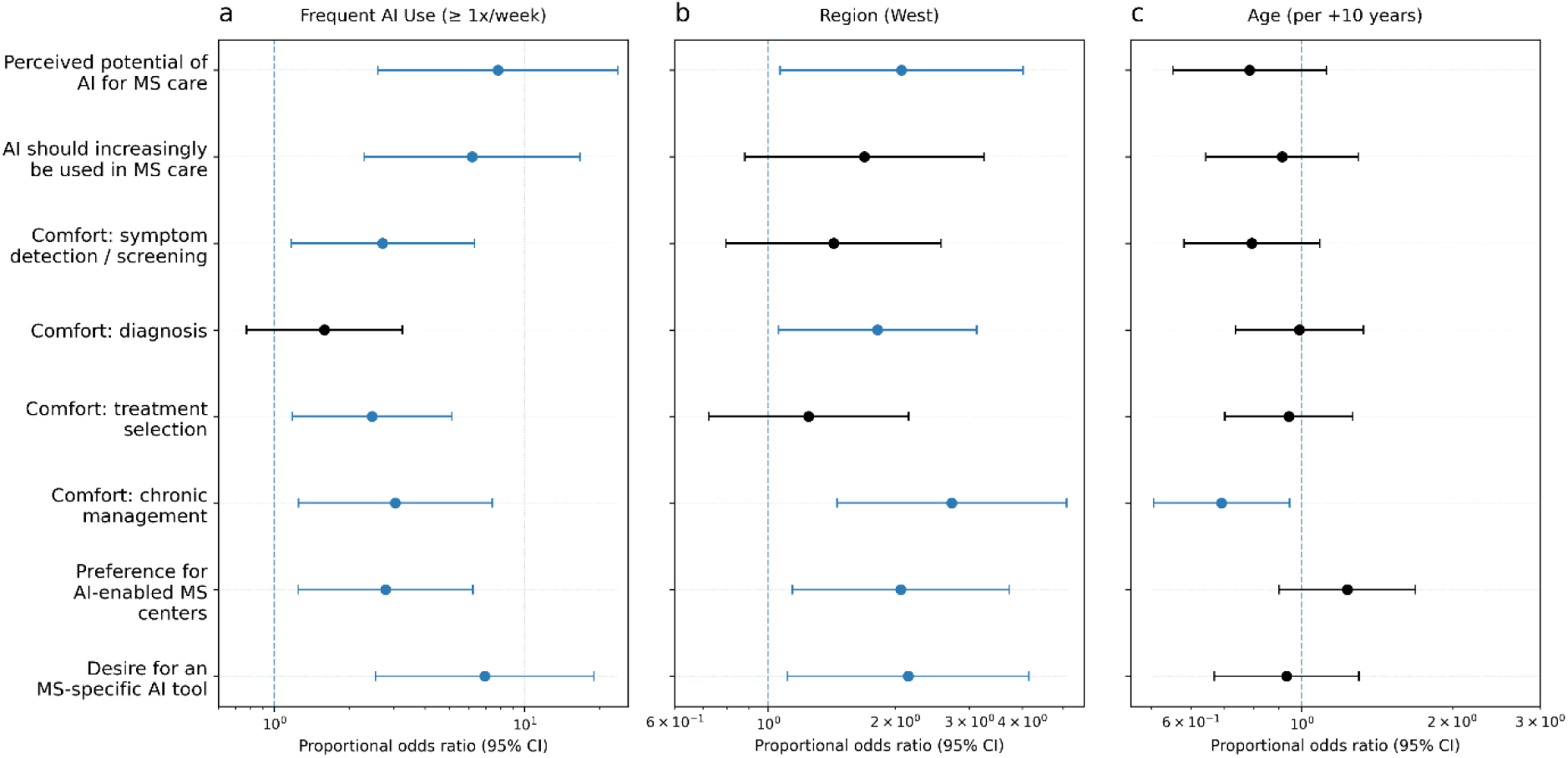
Predictors of artificial intelligence acceptance across eight clinical domains among people with MS: impact of frequent general AI use, region, and age. Forest plots show adjusted proportional odds ratios (POR; log scale) with 95% confidence intervals for (a) frequent general artificial-intelligence use (≥1x/week vs <1x/week), (b) region (Western vs Eastern Germany), and (c) age (per +10 years), across the eight secondary outcomes. POR>1 indicates higher odds of endorsing a higher (more accepting) response category. Estimates come from proportional-odds (ordinal logistic, logit link) models with outcomes collapsed to three levels (negative: 1–2; neutral: 3; positive: 4–5). All effects are adjusted for further covariates (demographics, disease characteristics, and artificial-intelligence literacy groupings). AI: Artificial Intelligence; MS: Multiple Sclerosis.

## Discussion

In this cross-sectional survey of pwMS, we found moderate overall acceptance of AI in MS care, but acceptance was highly use-case dependent. Two patterns stood out: a pronounced “responsibility gradient,” with greater comfort for supportive applications (e.g., screening, chronic management) and lower comfort for high-stakes tasks (diagnosis, treatment selection), and experiential familiarity—operationalized as frequent general AI use —emerged as the strongest and most consistent predictor of acceptance, whereas clinical indicators such as disability and disease duration were not independently associated.

Interestingly, our results suggest that patient acceptance is shaped less by disease burden than by digital inclusion and everyday exposure to AI. While a recent lager multinational study of stationary patients found that poorer health status was a significant predictor of lower AI acceptance, we observed no such association with disability in our MS cohort (5).

The strong association between frequent general AI use and both the composite score and multiple single-item outcomes is consistent with the Technology Acceptance Model. This framework proposes that perceived usefulness and perceived ease of use are the primary drivers of adoption, both of which are likely enhanced by the patients’ prior experience and familiarity with the technology.(17). Importantly, prior work suggests pwMS often show strong health information engagement and limited information avoidance (18), indicating that reluctance toward AI may not stem from disinterest or low competence, but from uncertainty about how AI works, what it can and cannot do, and how it will be governed in clinical settings. This is clinically actionable: unlike disease severity, familiarity can be increased through guided, low-risk exposure and patient education (13, 19).

Acceptance was not uniform across clinical domains. PwMS were more comfortable with AI in supportive roles but more cautious when AI was framed as influencing high-stakes medical decisions. This aligns with evidence from other patient populations showing stronger acceptance of AI as an assistive tool than as an autonomous decision-maker (5, 20, 21). Moreover, the observed’responsibility gradient’ aligns with recent global findings suggesting that patient caution correlates directly with the personal stakes of the clinical application (5). Importantly, most respondents preferred a collaborative model where clinicians retain final responsibility. This preference should be treated as a design requirement rather than a barrier: AI tools in MS care are more likely to gain trust when implemented as clinician-support systems embedded in shared decision-making and accompanied by clear accountability structures (15, 22).

Regional and age-related differences further suggest that acceptance is shaped by context. Residency in Western Germany was associated with greater acceptance across several domains, which may reflect differences in digital infrastructure, exposure to health technologies, and trust environments described in German digital health literacy research (23, 24). Age showed a negative association, particularly for AI-supported chronic management, consistent with previous work indicating that older adults’ concerns often center on operational errors, loss of personal interaction, privacy/security, and unequal access to care (25–27). Together, these findings highlight a potential equity issue: groups that may benefit from monitoring and supportive AI tools could also be those with lower baseline comfort, underscoring the need for age-sensitive and region-sensitive implementation approaches.

Taken together, our findings argue against a “one-size-fits-all” rollout. A pragmatic approach would be to start with high-acceptance use cases (monitoring, screening, administrative support), where benefits are tangible and perceived risk is lower, and then iteratively expand toward more decision-critical applications as familiarity and trust grow. Because familiarity appears to be a key lever, implementation strategies could include low-stakes onboarding (e.g., scheduling/administrative chatbots), transparent communication about data use and limitations, and patient-facing explanations that emphasize augmentation rather than replacement of clinician judgement (5, 15, 22, 28).This staged strategy may be particularly relevant for older pwMS and for settings with lower baseline acceptance (29, 30). Given the growing evidence for potential of assistive AI system in the care of MS, this is an urgent matter that requires both good clinical judgements as well as roll-out strategies that are coordinated with pwMS.

This study provides, to our knowledge, one of the first empirical baselines of AI acceptance in pwMS across multiple clinical domains, using both a reliable composite measure and domain-specific outcomes. However, several limitations should be considered. The cross-sectional design precludes causal inference. Recruitment through academic centres and digital channels likely introduced selection bias toward digitally engaged and highly educated participants. although MS serves as a valuable model for data-driven chronic care, these findings may have limited generalizability to populations with lower educational attainment, different disease phenotypes, or less access to specialized care. Additionally, MS diagnosis and disability were self-reported and not clinically verified. Collapsing Likert responses improved model stability but reduced response granularity. Finally, attitudes were assessed during a period of intense public discourse on AI (including large language models), which may have influenced responses and may not fully reflect stable long-term views.

Ultimately, our results suggest that the successful integration of AI into MS care is not merely a technological challenge, but a sociotechnical one. While the potential for algorithmic precision and newer models is clear, our findings suggest that perceived clinical utility is closely linked to clear communication about the tool’s role and to users’ prior familiarity with AI-enabled technologies, adding granularity by showing that caution is not a blanket rejection. Moving forward, the priority must shift from simply validating model performance to designing’human-in-the-loop’ workflows that respect the patient’s clear preference for augmented rather than automated care. By treating patient familiarity as a modifiable factor—cultivated through active education and continuous, low-risk engagement—clinicians and developers can bridge the current trust gap, ensuring that AI evolves from a theoretical asset into a clinically integrated partner in the long-term management of multiple sclerosis (13).

## Conclusion

PwMS appear open to AI especially for supportive functions while clearly preferring human accountability for high-stakes decisions. Our results highlight that acceptance is driven by experiential familiarity rather than clinical severity, a finding likely applicable to other data-rich chronic diseases. This suggests that careful, transparent, and staged implementation focused on building digital trust may be central to successful integration of AI into routine clinical practice.

## Materials and Methods

### Study design and setting

We conducted a cross-sectional, observational survey in Germany between Decembre 2024 and April 2025 using a structured, self-administered web-based questionnaire. Participants were recruited via digital channels (centre website, social media, newsletter) and during routine outpatient visits. Recruitment and distribution were facilitated by clinical staff at two academic MS centres (University Hospital Carl Gustav Carus Dresden; Medical Faculty and University Hospital Düsseldorf). Eligible participants were adults (≥18 years) with self-reported multiple sclerosis (MS; any subtype).

### Questionnaire development

As no suitable standard instrument existed for assessing patient attitudes toward AI in this specific context, a dedicated questionnaire was developed informed by previously published instruments tailored to the MS context (see Supplementary Material 1) (5, 31). The survey design was informed by the Technology Acceptance Model (TAM) but adapted to capture the specific nuances of trust in automated clinical decision-making. To ensure content validity, the item pool was generated and refined by a multidisciplinary board comprising neurologists, digital health scientists, and patient advocates.

The survey was hosted digitally on a secure platform on servers in Dresden. An initial information page described the study purpose, voluntariness, and data handling. No directly identifying information (e.g., names, contact details, IP addresses) was collected. Participation was voluntary; continuing to the questionnaire and submitting responses constituted implied consent. In line with local institutional policy for anonymous, minimal-risk survey research without collection of personally identifiable information, formal ethics committee approval and written informed consent were not required. The full questionnaire is provided in Supplementary Material 1.

### Survey instrument and measures

Items were developed to capture four domains:

1. Participant characteristics. Demographics (age, gender, education) and MS-related variables (disease course, year of diagnosis, and disability). Disability was self-reported based on a modified version of the Patient-Determined Disease Steps (PDDS) (32).
2. Technology access and use: ownership and use of devices (computer/laptop, smartphone/tablet, basic mobile phone, wearable), usage frequency, and health-related internet activities (e.g., MS information seeking; communication with clinicians/peers).
3. AI literacy and exposure: self-rated AI knowledge and frequency of general and health-related AI use (including health-related queries).
4. Attitudes and preferences regarding AI in MS care: perceived potential of AI, comfort with AI-supported care, and preferences for decision-making responsibility. Acceptance was assessed across eight predefined clinical domains.

Data were collected using LimeSurvey. No third-party data sharing occurred. Raw exports were screened for completeness and plausibility prior to analysis.

### Statistical analysis

Continuous and ordinal variables are reported as mean (SD) or median [IQR], and categorical variables as n (%). Likert outcomes were analysed at the item level using distributional summaries. Disease duration was calculated as survey year minus year of diagnosis. Associations between age, disease duration, and disability (PDDS) were assessed using Spearman’s rank correlation coefficients (r_s_). For subgroup analyses, participants were dichotomised using median splits for age (51 years) and disease duration (11 years). Regional location (Eastern vs Western Germany) was derived from the first three digits of the self-reported postal code. PDDS was grouped as low (0–2) versus high (3–8) following prior work (32, 33). Education was categorized as low (non-tertiary education: including vocational training and university entrance qualifications) vs high (tertiary: including degree from a university or university of applied sciences) adapted from the international standard classification of education (34). Device and internet use frequencies were dichotomised as ≥1/day versus <1/day.

AI literacy groupings were defined from three items: perceived AI knowledge (none/few vs good/expert), general AI use frequency (≥1×/week vs <1×/week), and health-related AI use frequency (≥1×/week vs <1×/week). Furthermore, an “AI attitudes composite” was formed based on AI-preference outcomes, internal consistency was evaluated with Cronbach’s alpha (higher scores = greater acceptance). Between-group comparisons in AI-preference outcomes used Mann-Whitney U tests on Likert-scale scores and an effect size (*r*) was computed (35). Differences in acceptance rates across the eight clinical domains were tested using Cochran’s Q with Bonferroni-corrected pairwise comparisons. Predictors of AI acceptance were examined using multivariable linear regression for the AI attitudes composite (primary outcome) and multivariable proportional-odds (ordinal logistic, logit link) models for each AI-preference item (secondary outcomes). To stabilise estimates and improve interpretability, Likert outcomes were collapsed into three levels: negative (1–2), neutral (3), and positive (4–5). All models included demographic variables, disease-related variables, and AI literacy groupings as predictors. Proportional odds ratios with 95% confidence intervals were reported for ordinal models. Model assumptions were checked; heteroscedasticity-robust standard errors were used. Predictors with near-zero variance (e.g., daily smartphone or internet use) were excluded.

Analyses were performed in IBM SPSS Statistics (v29; IBM, Somers, NY, USA). Two-sided P < 0.05 was considered statistically significant. Figures were generated in Python 3 (Matplotlib v3.10.0; Seaborn v0.13.2; NumPy v2.0.2).

## Data Availability

The datasets generated and/or analysed during the current study are available in the Open Science Framework (OSF) repository: https://osf.io/phqwg/overview?view_only=e3a0dfe190d3471d8e053f3323e1afcb.

## Contributions

H.I. and T.Z. conceived and designed the study. H.I., I.V., and R.H. developed the survey methodology and instrument. H.I., L.M., N.H., I.V., I.G.-C., and M.P. were responsible for data acquisition and curation. H.I., D.S. and R.H. performed the formal statistical analysis and visualization. H.I. wrote the original draft of the manuscript. L.M., J.W., S.M., H.B.H., S.G., and M.P. provided critical revision of the manuscript for important intellectual content. T.Z. supervised the project. All authors read and approved the final version of the manuscript.

## Acknowledgments

The authors would like to thank all people with MS who participated in this study for their time and valuable insights. Special thanks to aMStart and the MS Perspektive podcast for their assistance with study recruitment and dissemination. This work was funded by the German Research Foundation (DFG, Deutsche Forschungsgemeinschaft) as part of Germany’s Excellence Strategy – EXC 2050/2 – Project ID 390696704 – Cluster of Excellence “Centre for Tactile Internet with Human-in-the-Loop” (CeTI) of Technische Universität Dresden.

